# Impact of sarcopenic obesity on post-hepatectomy bile leakage for hepatocellular carcinoma

**DOI:** 10.1101/2023.05.15.23290011

**Authors:** Hikaru Hayashi, Akira Shimizu, Koji Kubota, Tsuyoshi Notake, Hitoshi Masuo, Takahiro Yoshizawa, Kiyotaka Hosoda, Hiroki Sakai, Koya Yasukawa, Yuji Soejima

## Abstract

**Background:** Post-hepatectomy bile leakage (PHBL) is a potentially fatal complication that can arise after hepatectomy. Previous studies have identified obesity as a risk factor for PHBL. Therefore, we investigated the impact of sarcopenic obesity on PHBL in hepatocellular carcinoma (HCC) patients.

**Methods:** In total, we enrolled 409 patients who underwent hepatectomy without bilioenteric anastomosis for HCC between January 2010 and August 2021. Patients were grouped according to the presence or absence of PHBL. Patient characteristics including body mass index and sarcopenic obesity were then analyzed for predictive factors for PHBL.

**Results:** Among the 409 HCC patients included in this study, 39 developed PHBL. Male gender, hypertension and cardiac disease, white blood cell counts, the psoas muscle area and visceral fat area, and intraoperative blood loss were significantly increased in the PHBL (+) group compared with the PHBL (−) group. Multivariate analysis showed that independent risk factors for the occurrence of PHBL were intraoperative blood loss ≥370 mL and sarcopenic obesity.

**Conclusion:** Our results show that it is important to understand whether a patient is at high risk for PHBL prior to surgery and especially to reduce intraoperative blood loss during surgery for patients with risk factors for PHBL.

## Introduction

Post-hepatectomy bile leakage (PHBL) is one of the most common and notable complications after hepatectomy, occurring in approximately 5% of patients after hepatectomy,[1, 2] and can lead to surgical site infection (SSI) or post-hepatectomy liver failure (PHLF).[3] According to previous reports, factors such as liver cirrhosis, non-anatomical hepatectomy, and obesity have been reported to be risk factors for PHBL.[4] Recently, sarcopenic obesity has been reported to be associated with postoperative outcomes for various carcinomas. Kim et al.[5] showed that sarcopenic obesity was an independent risk factor for increased mortality in patients with gastric cancer. Furthermore, Kobayashi et al.[6] reported that sarcopenic obesity was a significant prognostic factor for poor overall survival and relapse-free survival after hepatectomy for hepatocellular carcinoma (HCC). However, few studies have reported the relationship between short-term outcomes, especially PHBL, and sarcopenic obesity after hepatectomy for HCC. The purpose of this study was to evaluate the impact of sarcopenic obesity on the occurrence of PHBL after hepatectomy for HCC.

## Material and methods

In total, 409 patients who underwent hepatectomy without bilioenteric anastomosis for HCC between January 2010 and August 2021 were enrolled in this study. Patients were categorized into the following two groups on the basis of the presence or absence of PHBL: PHBL (+) and PHBL (−). Patient characteristics and relevant clinicopathological variables, surgical details, and short-term outcomes were recorded. This study was approved by the Biological and Medical Research Ethics Committee of Shinshu University School of Medicine (approval no. 5701) and was conducted in accordance with the principles of the Declaration of Helsinki. Due to the retrospective property of this study and absence of invasive interventions, the requirement for written consent was waived by the review board, and consent was obtained through an opt-out method. The data was analyzed anonymously on the basis of medical records.

### Perioperative management

In this study, hepatectomies were performed by several surgeons. Parenchymal transection was performed using an ultrasonic dissector and/or the clamp-crushing technique. The intermittent Pringle maneuver (PM) was routinely used to control intraoperative blood loss for 15 min, followed by 5 min of reperfusion; this process was repeated as needed. Abdominal drains were routinely placed along the cut surface of the liver. Postoperatively, bilirubin concentrations in the drainage fluid were routinely measured twice per week for surveillance of bile leakage. Furthermore, for several days after surgery, the presence or absence of fluid collection around the site of hepatectomy was checked every day by ultrasonography. For PHBL, additional percutaneous drainage or exchanging drain tubes were performed as needed. The end of follow-up was December 2021 or the time of discharge.

### Definitions

Pathological findings were evaluated in accordance with the American Joint Committee on Cancer Staging Manual, 7th edition[7] (AJCC), and liver cirrhosis was defined as a fibrosis score of 4 using the new Inuyama classification[8]. Postoperative complications were graded using the Clavien–Dindo classification.[9] PHLF and PHBL were diagnosed and graded according to the criteria of the International Study Group of Liver Surgery.[10, 11] Major hepatectomy was defined as resection of three or more Couinaud’s segments of the liver. Anatomical resection included Couinaud’s segmentectomy, sectionectomy, hemihepatectomy, and trisectionectomy.

Multidetector-row computed tomography (CT) was performed within 4 weeks before surgery for diagnostic and staging purposes and was used to evaluate sarcopenia, which was defined in accordance with the international consensus[12] as a skeletal muscle index (SMI) of <52.4 cm^2^/m^2^ for men and <38.9 cm^2^/m^2^ for women. SMI was defined as the total muscle area measured on an axial section through the third lumbar vertebra (L3) when both pedicles were visible with a preestablished density threshold of −29 to +150 Hounsfield units normalized for stature (Fig 1). Similarly, visceral fat area (VFA) and subcutaneous fat area were also measured, and sarcopenic obesity was defined by the VFA/SMI ratio.[13, 14] The cut-off value for sarcopenic obesity was defined in accordance with the maximum sensitivities and specificities for predicting PHBL in receiver operating characteristic (ROC) curve analysis; the cut-off value was 1.65. CT scans were checked by two qualified physicians using a Synapse Vincent FN-7941 (Fujifilm, Tokyo, Japan).

**Fig 1.**
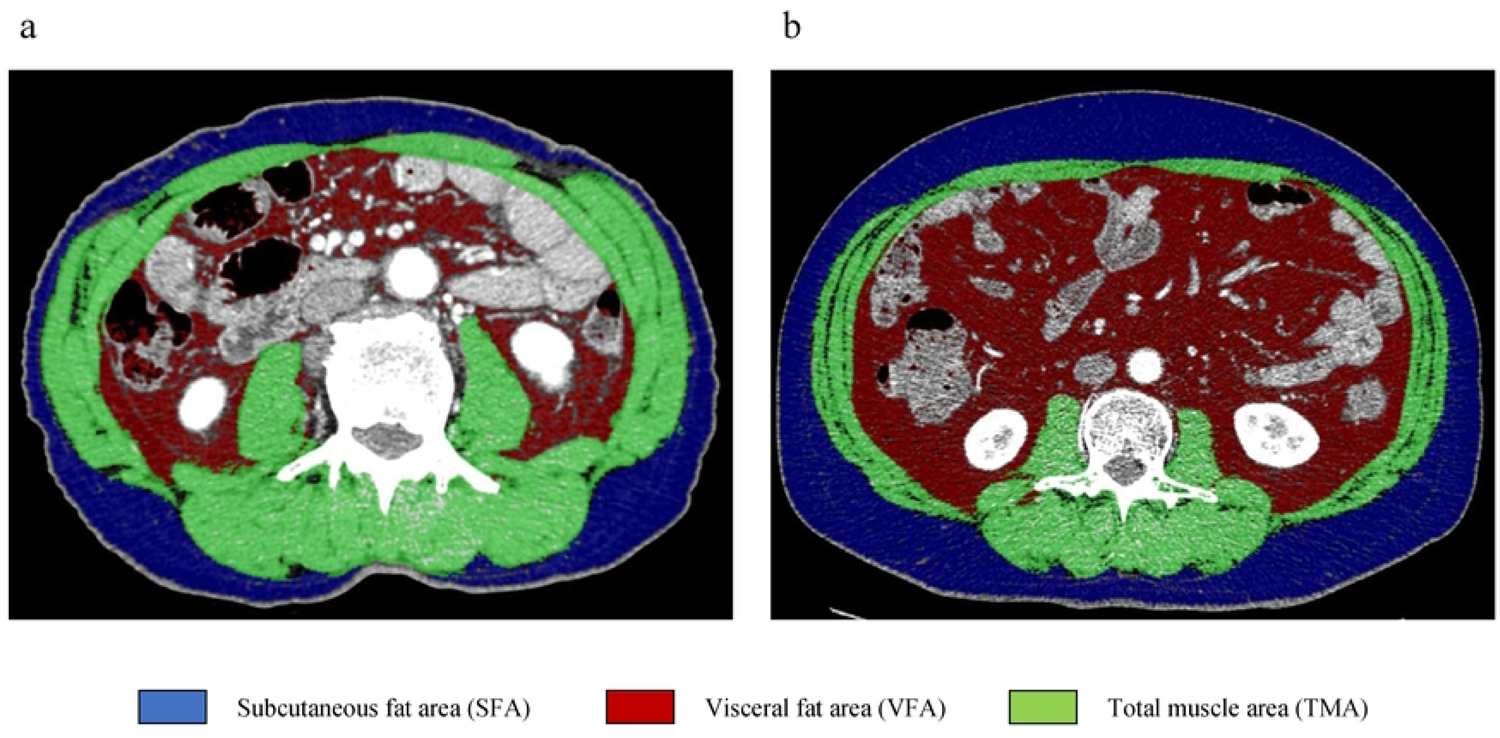
Computed tomography image analysis of a third lumbar vertebra. Green area, muscle; red area, visceral fat; blue area, subcutaneous fat. (a) A patient without sarcopenic obesity. (b) A patient with sarcopenic obesity.

### Statistical analysis

All data were collected by a research assistant and stored in a computer database. Statistical analysis was performed by the Chi-square test or Fisher’s exact test to compare categorical variables and by the Mann–Whitney U test to compare continuous variables. ROC curve analyses for the predictive parameters were used to evaluate associations with PHBL, with the Youden index used to determine cut-off values. Multivariate analysis using a logistic regression model was conducted to identify independent significant predictive factors for the occurrence of PHBL. All statistical analyses were performed using JMP Pro 16.2 (SAS Institute Inc., Cary, NC, USA).

## Results

### Patient characteristics

During the study period, a total of 409 patients underwent hepatectomy. Among all patients, 39 (9.5%) developed PHBL. A comparison of the patients’ background characteristics and preoperative comorbidities according to the presence or absence of PHBL is summarized in Table 1. The ratio of male gender and the prevalence of hypertension and cardiac disease were higher in the PHBL (+) group than in the PHBL (−) group (89.7% vs. 74.3%; 74.4% vs. 53.0%; 28.2% vs. 13.0%; *P* = 0.020, 0.009, and 0.018, respectively). The indocyanine green retention rate at 15 min was lower in the PHBL (+) group than in the PHBL (−) group (9.7% vs. 12.0%; *P* = 0.026). There were no significant differences between the two groups regarding other variables, including age, etiology, or Child–Pugh classification.

### Preoperative muscle and fat areas

A comparison between the two groups regarding muscle and fat areas is shown in Table 1. At the L3 level, the psoas muscle area and VFA were significantly larger in the PHBL (+) group than in the PHBL (−) group (16.6 vs. 14.0 cm^2^; 135.3 vs. 96.8 cm^2^; *P* = 0.016 and 0.003, respectively). In contrast, SMI was comparable between the two groups. Although the ratio of patients with sarcopenia was comparable, the ratio of patients with sarcopenic obesity was significantly higher in the PHBL (+) group than in the PHBL (−) group (89.7 vs. 59.7%; *P* < 0.001).

### Surgical outcomes

Among the entire cohort, the median surgical duration time was 340 min (range: 82–990 min), the median inflow occlusion time was 60 min (range: 0–247 min), and the median intraoperative blood loss was 300 mL (range: 0–5500 mL). The surgical outcomes of each group are detailed in Table 1. No significant differences were observed between the two groups with regards to the type of hepatectomy, the ratio of primary hepatectomy, or anatomical hepatectomy. Additionally, there was no significant difference in the ratio of medial sectionectomy or central bisectionectomy. Furthermore, surgical duration and inflow occlusion times were comparable between the two groups. However, intraoperative blood loss was greater in the PHBL (+) group than in the PHBL (−) group (430 vs. 278 mL, *P* = 0.002). As a result, the percentage of intraoperative blood transfusions tended to be higher in the PHBL (+) group (25.6 vs. 13.8%; *P* = 0.065).

### Histopathological findings

Histopathological features are summarized in Table 2. The prevalence of liver cirrhosis was comparable between the two groups (*P* = 0.074). In contrast, the ratio of multiple tumors tended to be lower in the PHBL (+) group than in the PHBL (−) group (15.4 vs. 27.0, *P* = 0.097). Furthermore, the pathological AJCC staging tended to be lower in the PHBL (+) group (*P* = 0.071). There were no significant differences between the two groups in maximum tumor diameter, the ratio of microvascular invasion, or the R0 resection rate.

### Short-term outcomes

Regarding short-term outcomes (Table 3), 225 patients (55.0%) developed postoperative complications, including 70 (17.1%) with complications of grade III or higher.[9] One patient (0.2%) among the entire cohort died due to grade C PHLF. In the PHBL (+) group, 12 patients (30.8%) developed grade B PHBL and 27 (69.2%) developed grade A PHBL, while no patients developed grade C PHBL. As for PHLF, all cases of PHLF in the PHBL group were grade A, and there was no significant difference between the two groups in the occurrence of PHLF. Moreover, incidences of intra-abdominal infection or pleural effusion were comparable between the two groups. However, postoperative hospital stays were longer in the PHBL (+) group (14 vs. 12 d; *P* = 0.036).

### Risk factors for PHBL

The cut-off value for intraoperative blood loss was defined using ROC curve analysis, as well as sarcopenic obesity. The independent risk factors for the occurrence of PHBL were intraoperative blood loss ≥370 mL (odds ratio [OR]: 2.25; 95% confidence interval [CI]: 1.10–4.59; *P* = 0.026) and sarcopenic obesity (OR: 4.08; 95% CI: 1.37– 12.1; *P* = 0.011) (Table 4). Furthermore, if the analysis was limited to only patients with primary hepatectomy, sarcopenic obesity (OR: 3.61; 95% CI: 1.21–10.8; *P* = 0.022) was the only independent risk factor for the occurrence of PHBL (data not shown).

A scatter diagram of the correlation between intraoperative blood loss and sarcopenic obesity (VFA/SMI level) was created (Fig 2). Spearman’s rank correlation analysis showed low correlation between the amount of intraoperative blood loss and the VFA/SMI value (*ρ* = 0.201; *P* < 0.001). No patients with intraoperative blood loss <370 mL and without sarcopenic obesity developed PHBL (0 of 108 patients). In contrast, patients with intraoperative blood loss ≥370 mL and sarcopenic obesity had the highest ratio of the occurrence of PHBL among all combinations of these two parameters (17.8%, 21 of 118 patients). Additionally, ROC curve analysis of combined intraoperative blood loss and sarcopenic obesity had a higher area under the curve value (0.708; *P* = 0.013) than either parameter alone (Fig 3).

**Fig 2.**
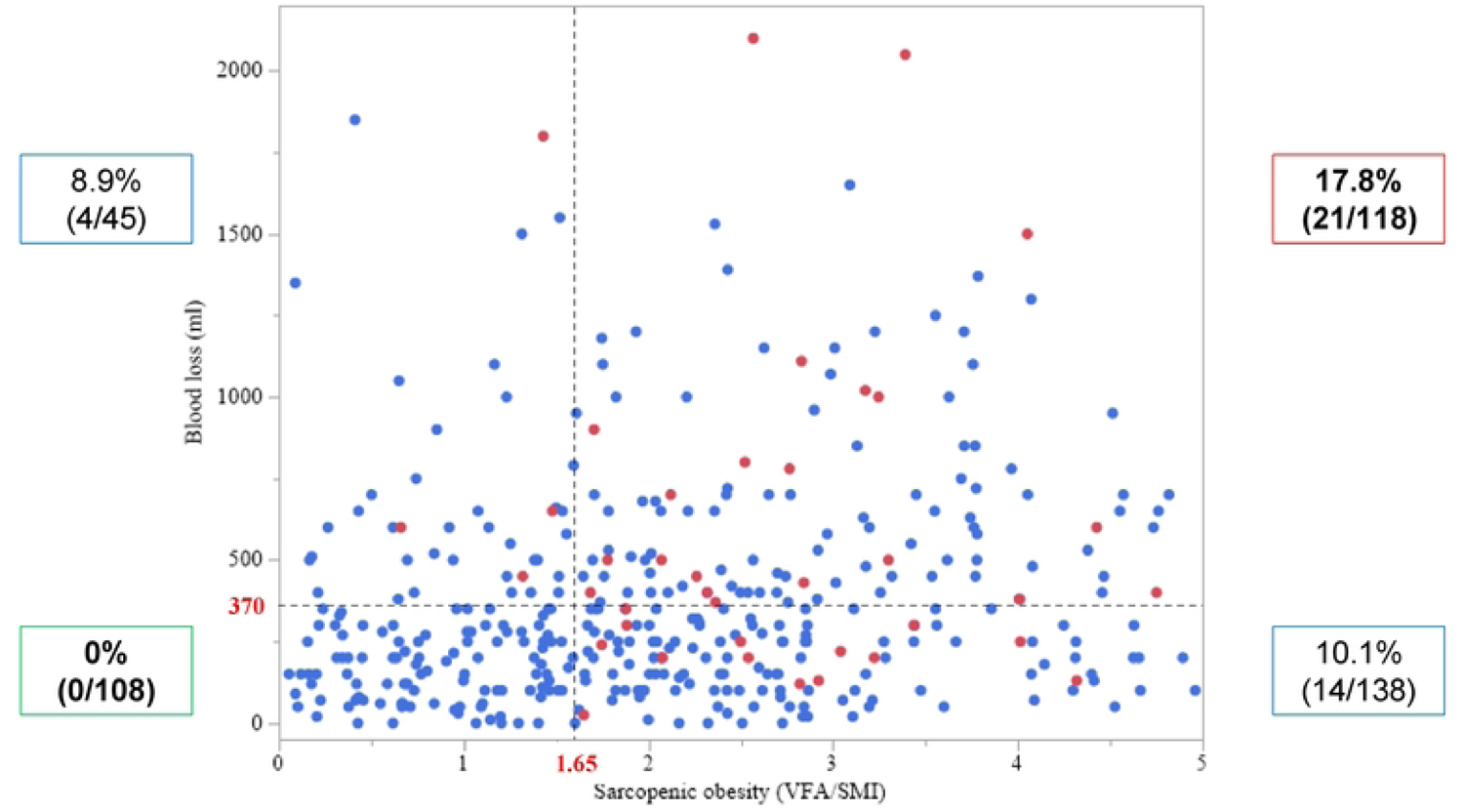
Scatter diagram of the correlation between visceral fat area (VFA)/skeletal muscle index (SMI) and intraoperative blood loss. Coefficients (ρ) and *P*-values were calculated using the Spearman’s rank correlation analysis. Red circles indicate patients with post-hepatectomy bile leakage (PHBL), and blue circles indicate patients without PHBL.

**Fig 3.**
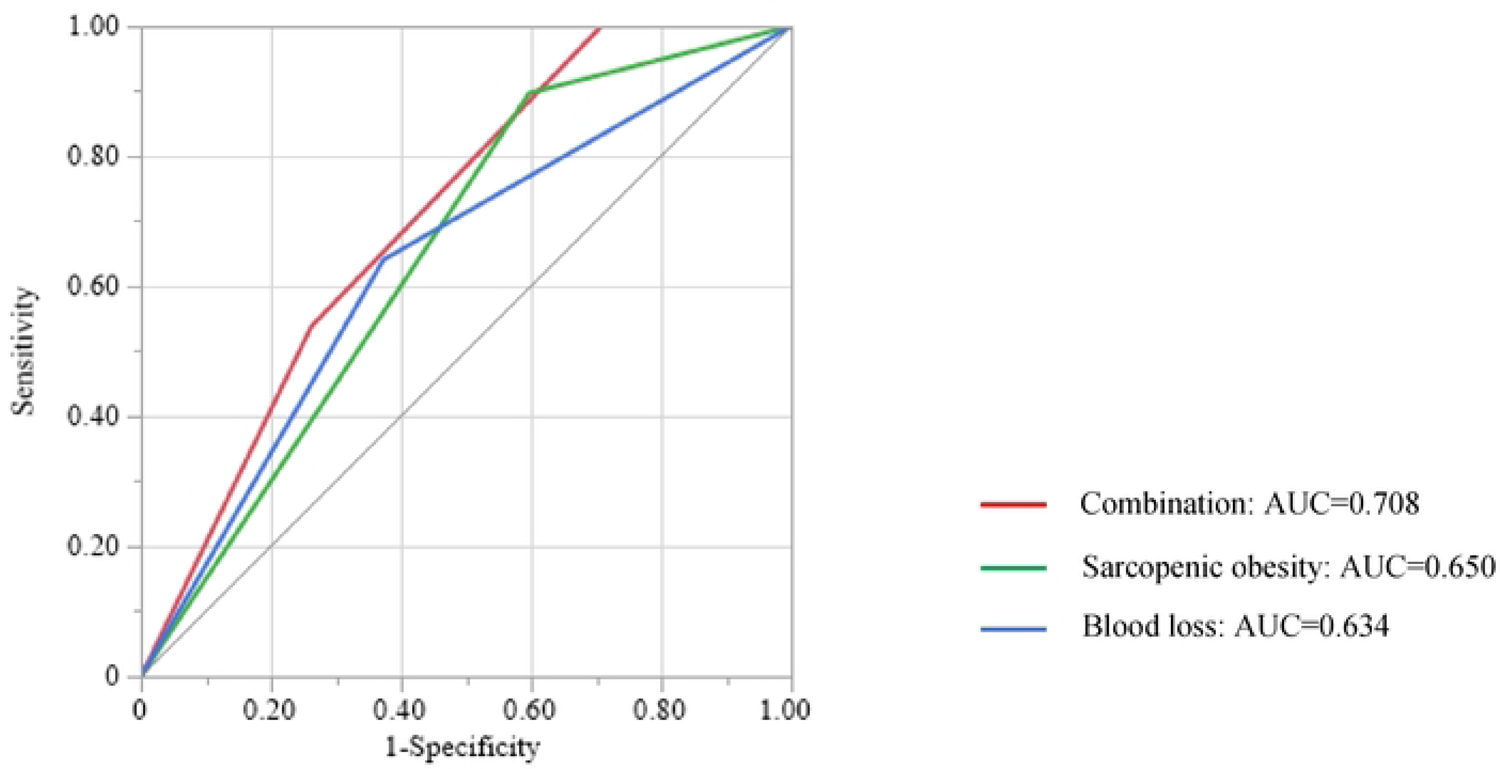
Receiver operating characteristic (ROC) curve analysis for the occurrence of PHBL. Red, ROC curve of the combination of sarcopenic obesity and intraoperative blood loss; green, ROC curve of sarcopenic obesity; blue, ROC curve of intraoperative blood loss.

## Discussion

Recently, the target of hepatectomy has been expanding, for example, for liver metastasis of various carcinomas; hepatectomy has become of increasing importance despite advances in drug therapy. PHBL is one of the most common postoperative complications after hepatectomy. Although most PHBL can be cured by conservative treatment, it can lead to PHLF or sepsis from an infection. Shehta et al.[15] reported that PHBL occurred in 5.8% of hepatectomy patients, and the PHBL (+) group had higher grades of PHLF. Furthermore, Lo et al.[16] showed that biliary complications including PHBL developed in 8.1% of hepatectomy patients and that these complications carried high risks for PHLF and mortality. In contrast, Okabayashi et al.[17] revealed that high body mass index (BMI), high intraoperative blood loss, presence of PHBL, and poor postoperative glucose control were risk factors for the occurrence of SSI from a multivariate analysis. In the present study, no significant differences were observed in the occurrence of PHLF and SSI between the PHBL (+) and PHBL (−) groups. This might be attributed to close monitoring and early intervention and appropriate usage of prophylactic antibiotics at our institute. The results of many previous studies indicate that PHBL itself is associated with other postoperative complications after hepatectomy; hence, it is desirable and reasonable to try to identify, before surgery, the population who are at a high risk for PHBL, to improve postoperative outcomes.

Obesity is one of the main health concerns that needs to be solved, not only in Japan, but also particularly in developed countries. Obesity is clearly related to lifestyle diseases, such as hypertension and cardiovascular diseases, HCC from nonalcoholic steatohepatitis and colorectal cancer.[18] For example, a large database retrospective study[19] showed that obesity was an independent risk factor for colorectal cancer across all age groups compared with the general population. BMI is the most common and simple indicator of obesity. Almost all previous reports that have shown a relationship between obesity and postoperative outcomes have been based on BMI. However, BMI can be inaccurate at times because it depends on muscle mass as well as fat mass. Therefore, we focused on sarcopenic obesity as a more objective and accurate indicator. Sarcopenic obesity was an independent risk factor for the occurrence of PHBL in this study, whereas BMI was not.

Sarcopenic obesity is a condition in which lean body mass is lost while fat mass is preserved or even increased and is one of the indicators for patient nutritional assessment.[20] Previous studies have suggested that sarcopenic obesity is a risk factor for short- and long-term outcomes after surgery. For example, Runkel et al.[21] reported that sarcopenic obesity was an independent risk factor for overall complications after hepatectomy for colorectal liver metastases. As for the association of fat and PHBL, our previous study[14] reported that sarcopenic obesity was an independent risk factor for the occurrence of postoperative pancreatic fistula after pancreaticoduodenectomy. The underlying reason was considered to be that the adipose tissue around the pancreatic duct might complicate anastomosis, decrease local blood flow for wound healing, and produce inflammatory cytokines that impede healing. Inflammatory cytokines produced by excess adipose tissue can cause delayed biliary wound healing, leading to PHBL. Therefore, the results of this study fit with existing data.

There are also several reports on the relationship between intraoperative bleeding and PHBL. Wang et al.[22] reported that tumor size, type of tumor, surgical duration time, blood loss, and blood transfusion were the independent risk factors for PHBL. Intraoperative blood loss might cause liver damage and reduced blood flow around the bile duct, resulting in bile leakage. Another possibility is that surgery with a wide surface area for the incision exposes more bile ducts at the surface.[23] Hepatectomy with such a wide surface area of the incision may increase intraoperative blood loss, and this study might reflect this possibility. However, this study was a retrospective study, and the area of the cut surface could not be measured accurately. Furthermore, as mentioned in the results, no significant difference was observed in the type of hepatectomy, such as medial sectionectomy or central bisectionectomy.

Occluding the hepatic inflow pedicle, also known as the Pringle maneuver (PM), is a widely accepted method for reducing intraoperative blood loss. However, the PM results in ischemia-reperfusion changes.[24] According to several animal studies, ischemia-reperfusion injury owing to hilar vascular clamping can accelerate tumor growth, stimulate tumor cell adhesion, and promote metastasis.[25] Furthermore, previous reports have shown that PM duration for HCC was associated with postoperative long-term outcomes.[26, 27] However, because of recent advances in surgical techniques, PM is not needed in every operation. Maurer et al.[28] reported that major resection without PM is feasible and safe and might reduce liver damage and failure. However, deciding to perform PM during hepatectomy should be based on risk assessment and operative difficulties. The appropriate intraoperative supportive techniques to complete the scheduled operation with minimal intraoperative blood loss are important.

This study had several limitations. First, this was a single-center retrospective study, and selection bias is possible. Second, the cut-off values for sarcopenic obesity and intraoperative blood loss were defined in accordance with the maximum sensitivities and specificities for predicting PHBL from ROC curve analysis. Therefore, this cut-off value might not be applicable to other institutions. Despite these drawbacks, we believe that our findings are of interest to surgeons because, to the best of our knowledge, this is the first study to investigate the association between sarcopenic obesity and PHBL in HCC patients.

## Conclusion

Sarcopenic obesity and intraoperative blood loss were significant risk factors for the occurrence of PHBL. It is important to preoperatively understand whether a patient is at high or low risk for PHBL for early therapeutic intervention.

## Data Availability

Data cannot be shared publicly because of including personal information. Data are available from the Biological and Medical Research Ethics Committee of Shinshu University School of Medicine (contact via corresponding author) for researchers who meet the criteria for access to confidential data.

## Acknowledgments

We thank James P. Mahaffey, PhD, from Edanz (https://jp.edanz.com/ac) for editing a draft of this manuscript.

